# Why are people cosmetic skin whitening? A systematic review

**DOI:** 10.1101/2025.06.10.25329253

**Authors:** Simon N Williams, Trevor Webb, Anna-Leena Lohiniva, Elena Jardan, Lesley Onyon, Elena Altieri

## Abstract

**Importance:** Cosmetic skin whitening is a growing practice in a number of countries and long-term use has been shown to have potentially serious health risks. No reviews have documented the behavioral factors that help explain practices related to cosmetic skin whitening.

**Objective:** To explore the main behavioral (psychological and cognitive, social and cultural, and environmental) factors behind the practice of cosmetic skin whitening, and to explore which groups and characteristics are associated with the practice as well as what interventions have been designed to address this practice.

**Evidence Review:** Five databases (PubMed, Scopus, APA PsychINFO, ASSIA, and Web of Science’s Preprint Citation Index) were searched using adaptations of key search terms (e.g. “skin whitening”, “behavior”, “risk”). Studies were not restricted by date or country.

**Findings:** From 816 candidate studies, 43 studies were included in the final sample. Twelve main factors were identified: beauty; self-esteem; to attract a partner or get married; dissatisfaction with skin tone; low awareness of risks of skin whitening; to look whiter or fairer; for social status; social norms or social influence; for a job or to secure employment; colorism; the influence of advertising; and fashion. Prevalence of use ranged from 2-74%. Use was more common amongst women and younger adults but varied by country or region in terms of whether it was more commonly used by people with high or low formal education. Only one study documented an intervention designed to raise awareness of the harms of cosmetic skin whitening.

**Conclusions and Relevance:** Cosmetic skin whitening is a complex practice influenced by multiple behavioral factors. Findings should be used to inform theory-based interventions designed to reduce the prevalence of cosmetic skin whitening practices.

**Key points:** *Question:* What are the main behavioral factors behind the practice of cosmetic skin whitening, what demographic groups and characteristics are most likely to engage in skin whitening, and what interventions exist that are designed to reduce its prevalence.

*Findings:* Twelve main factors were identified: Beauty; self-esteem; to attract a partner or get married; dissatisfaction with skin tone; low awareness of risks of cosmetic skin whitening; to look whiter or fairer; for social status; social norms or social influence; for a job or to secure employment; colorism; the influence of advertising; and fashion.

*Meaning:* The practice of cosmetic skin whitening is a complex combination of psychological and cognitive, social and cultural and environmental factors. Themes identified can be used to inform tools for future research to understand the drivers of cosmetic skin whitening, as well as theory-based interventions designed to understand and address the practice of skin whitening.

---

Cosmetic skin whitening (‘skin bleaching’ or ‘skin lightening’) is the practice of using natural or synthetic substances (depigmenting agents) to lighten the skin tone or provide an even complexion. A variety of harmful agents including corticosteroids, fruit acids, hydroquinone and mercury are used which aim to either directly or indirectly reduce the melanin concentration in the skin.^1,2^ This practice is a matter of global public health concern.^3^ WHO has drawn attention to the dangers associated with the continued use of mercury in some skin-lightening products which is also in direct contravention of the Minamata Convention on Mercury.^4^ The International League of Dermatological Societies calls for global action as misuse of topical corticosteroids for skin bleaching grows.^5^ Long-term use of certain skin lightening products (SLPs) is linked to increased risk of skin cancer and conditions like scabies, high blood pressure, diabetes, neurological, and kidney problems.^6^

The practice of cosmetic (non-medical) skin whitening has a long history, dating back to antiquity,^7^ it has become more common in countries in Africa, Asia, the Middle East, and the Americas. Cosmetic skin whitening is linked to global racism, which is essentially discrimination based on skin color, and colorism, that is, the privileging of lighter skin within ethnic and racial groups.^7,8^ Although not all countries experiencing colorism have necessarily linked to colonialism, many countries where skin lightening have a colonial past.^7^ The current market for skin lightening products is estimated to be worth $8.8 billion annually and is projected to grow to $15.7 billion by 2030.^10^ A meta-analysis of 68 studies found that the lifetime (ever use) prevalence of skin whitening in the countries and populations studied was 27.67%.^11^ SLPs use has been found to be associated with those living in urban or semi-urban areas, younger adults (aged ≤30 years), and those with only primary school education.^11^ Although evidence generally suggests that use of SLPs is higher amongst females, research has also demonstrated relatively high use in males in studied countries and populations, indicating it is a unisex practice.^11^ There are multiple possible reasons for using SLPs for cosmetic purposes, including due to the perception of whiter skin being synonymous with status, health, youth, or beauty in some countries and cultures.^9,12^ The practice of cosmetic skin whitening needs to be understood in relation to historical factors, structural inequality and racial privilege.^13^ Colonial rule and slavery linked whiteness with modernity, civility, and social status embedding color hierarchies within local cultures.^1^ Additionally, in both colonial and non-colonial cultures (e.g. in some countries in Asia) darker skin has become equated with lower status occupations, for example outdoor laboring and farming where greater sun-exposure results in darker skin.^8^ ^13^

To the best of our knowledge, there are no systematic reviews on the reasons why people choose to use SLPs for non-medical, cosmetic reasons – specifically regarding the behavioral (i.e. psychological and cognitive, social and cultural, and environmental) factors of SLP use. A review of behavioral factors can help inform the design of targeted interventions which aim to reduce the public health harms of SLP use. This systematic review addresses the following key questions: (1) What are the main factors (including cognitive and psychological; social and cultural and environmental factors) behind the practice of skin whitening? (2) Which population groups and population characteristics are associated with the practice of skin whitening? (3) What interventions are designed to address the practice of skin whitening and what, if any, evidence is there of their effectiveness?

## Methods

### Search Strategy

A systematic review of the academic literature was performed, whereby candidate studies were retrieved from the following databases: PubMed, Scopus, APA PsychINFO, ASSIA, and Web of Science’s Preprint Citation Index (which searches across five major preprint repositories - arXiv, bioRxiv, medRxiv, chemRxiv, and Preprints.org.). The following generic search string was created and adapted to databases: ("skin lighten*" OR "skin whiten*" OR "Skin bleach*") AND (behavio* OR usage OR driver* OR reason* OR health* OR harm* OR risk*).

A specific date range was not set, and publication dates for retrieved candidate studies ranged from 1975-2024. Studies were not limited in terms of geographic scope. However, only studies in English were included. A full list of inclusion and exclusion criteria are provided in Box 1.

#### Box 1.

Inclusion and Exclusion criteria.

*Inclusion criteria*:

— English language studies

— Must contain empirical data (e.g. not commentary articles)

— Reporting on the non-medical use of skin whitening or bleaching products, specifically the problematic or misuse of such products for cosmetic reasons (e.g. not prescribed by a medical professional for the treatment of medical conditions such as melasma)

— contain data relating to the reasons, factors or drivers behind such use of skin whitening products.

*Exclusion criteria:*

— non-English language studies

— Articles not containing empirical data (e.g. commentary articles)

— Articles reporting on the medical use of skin whitening or bleaching products, specifically the problematic or misuse of such products for cosmetic reasons (e.g. not for the treatment of medical conditions such as melasma)

— Does not contain data relating to the reasons or factors behind use of such skin whitening products (e.g. focuses on different types and compositions of skin whitening products, focuses on types of health risks associated with skin whitening products, or generally discusses their use but not the reasons or factors behind such use)

Inclusion and exclusion criteria were determined by the research team (all listed authors) (see Box 1). All citations were screened independently by two reviewers (SW, TW) at the title and abstract level. Full papers were independently screened by two reviewers (SW, TW). Inter-rater reliability was assessed using Cohen’s Kappa (κ = 0.79), indicating substantial agreement, with remaining disagreements resolved by consensus discussions with a with a third reviewer (AL) (see PRISMA flow diagram). The coding framework for data extraction was developed by all members of the research team and was pilot tested by three authors (SW, TW, AL) on a small sample of papers. Data extraction was performed independently on all included papers by two members of the research team (SW, TW) with disagreements being resolved by consensus discussion which included a third reviewer (AL). The review research questions guided the thematic analysis, whereby key themes were drawn out across studies. In addition to key behavioral factors, key information about the study was extracted including details related to the study population and the type of study designs used - we applied The Oxford Centre for Evidence-Based Medicine (OCEBM) levels of evidence.^14^ Extracted data were reviewed, analyzed and grouped into themes inductively, with authors discussing by consensus what constitutes a significant theme - a combination of frequency and conceptual significance.^15^ Themes were subsequently written into this narrative synthesis.^15^

## Results

A total of 1,232 records were identified via the search strategy. Following removal of duplicates, 816 records were screened for eligibility using study titles and abstracts. The full texts of 48 studies were then assessed for eligibility. 43 papers met full inclusion criteria and were included in the final sample (see Figure 1, PRISMA flow diagram).

### Study characteristics

Overall, the 43 included studies used a variety of methods, with most (n=33) being cross-sectional quantitative surveys, but also including qualitative studies (n=3) and other or mixed-methods studies (n=5). Only two studies which met inclusion criteria documented an intervention designed to address the problematic practice of skin whitening. The included papers were global in scope, with studies from Africa (n=17); Asia (n=17); the Americas (n=7) and Europe (n=1). Most studies collected data from one individual country, with one study comparing across 26 countries. Table 1 summarizes the methods, study population and main findings of the studies included in this systematic review.

**Table 1:**
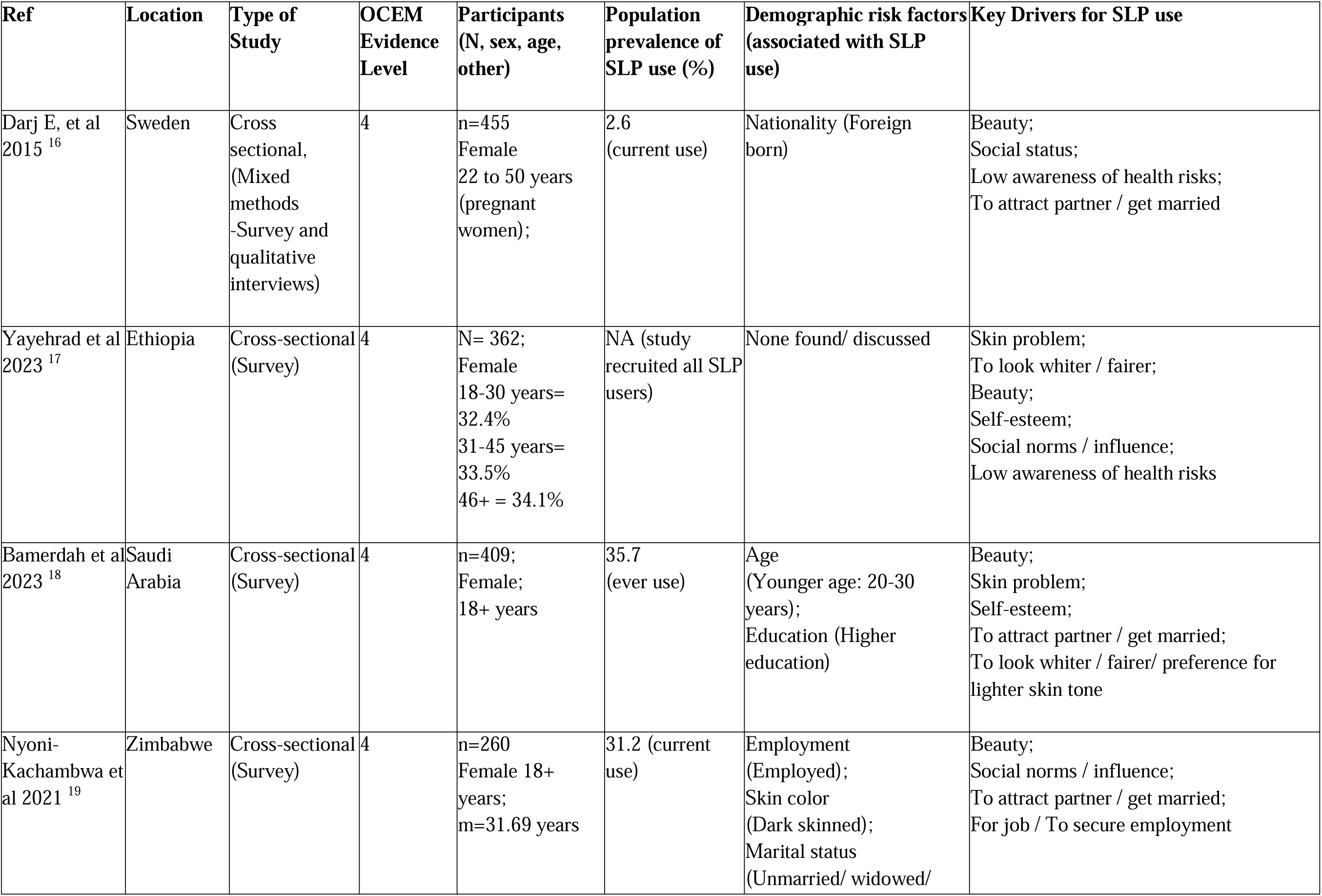
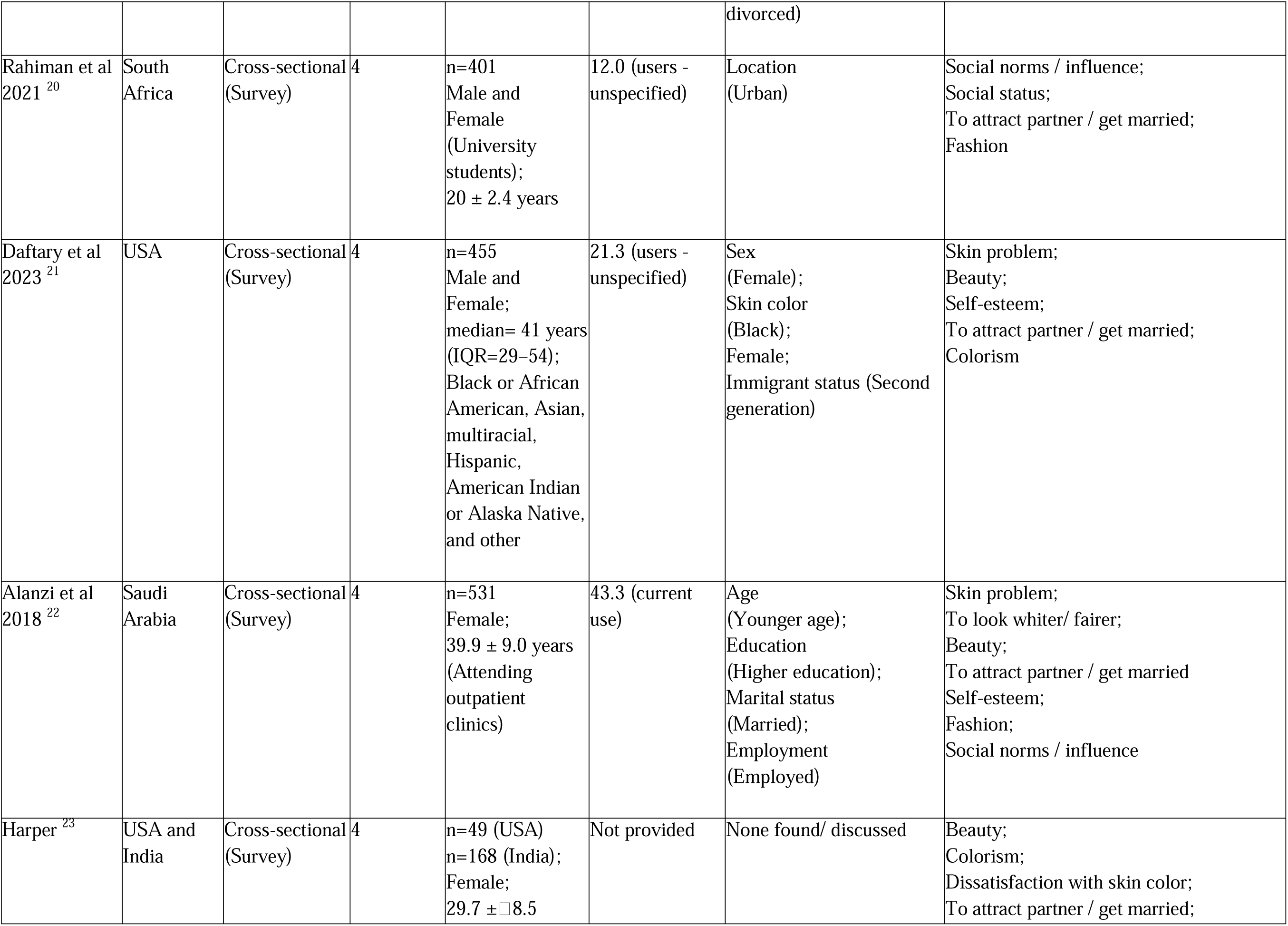
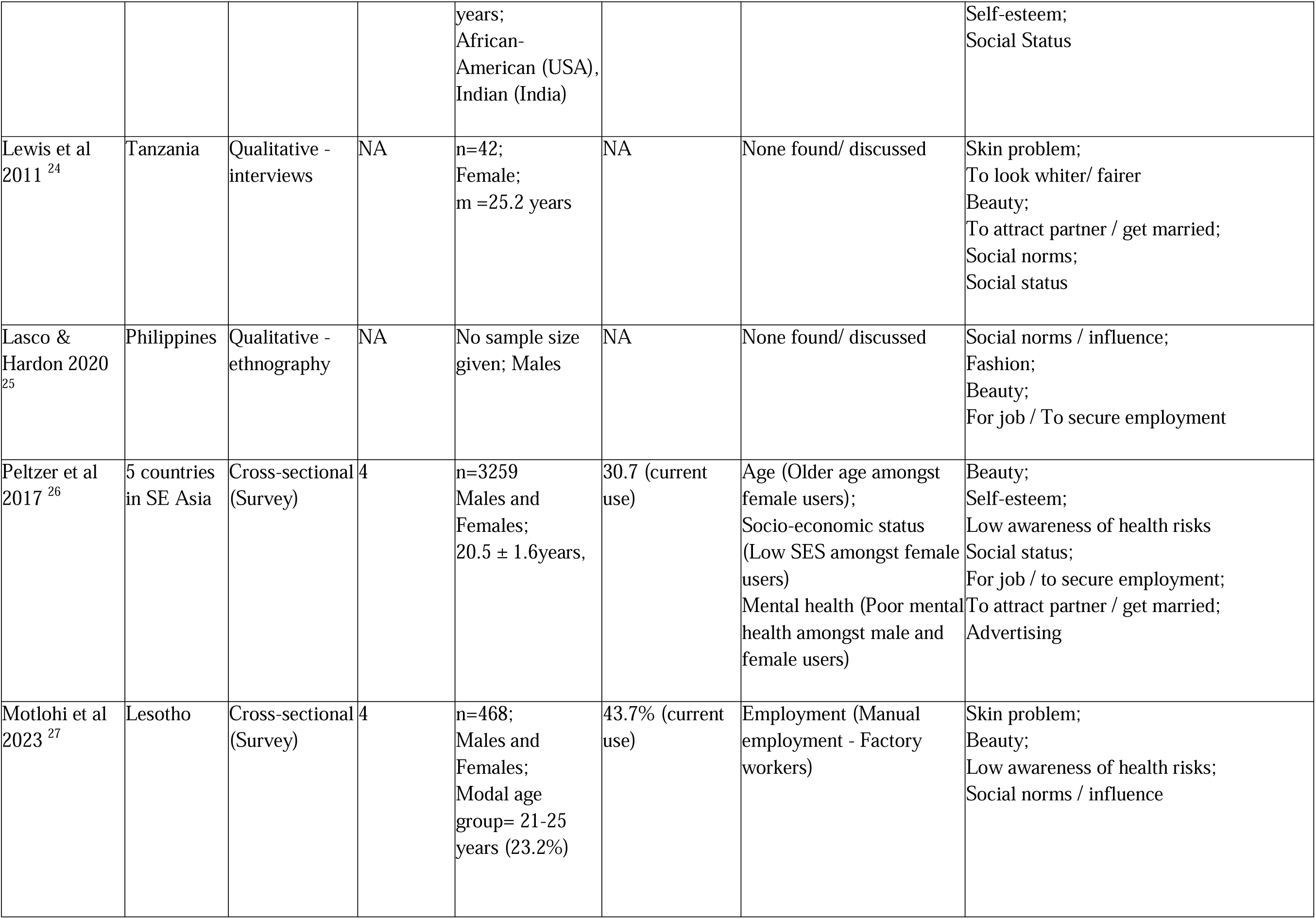
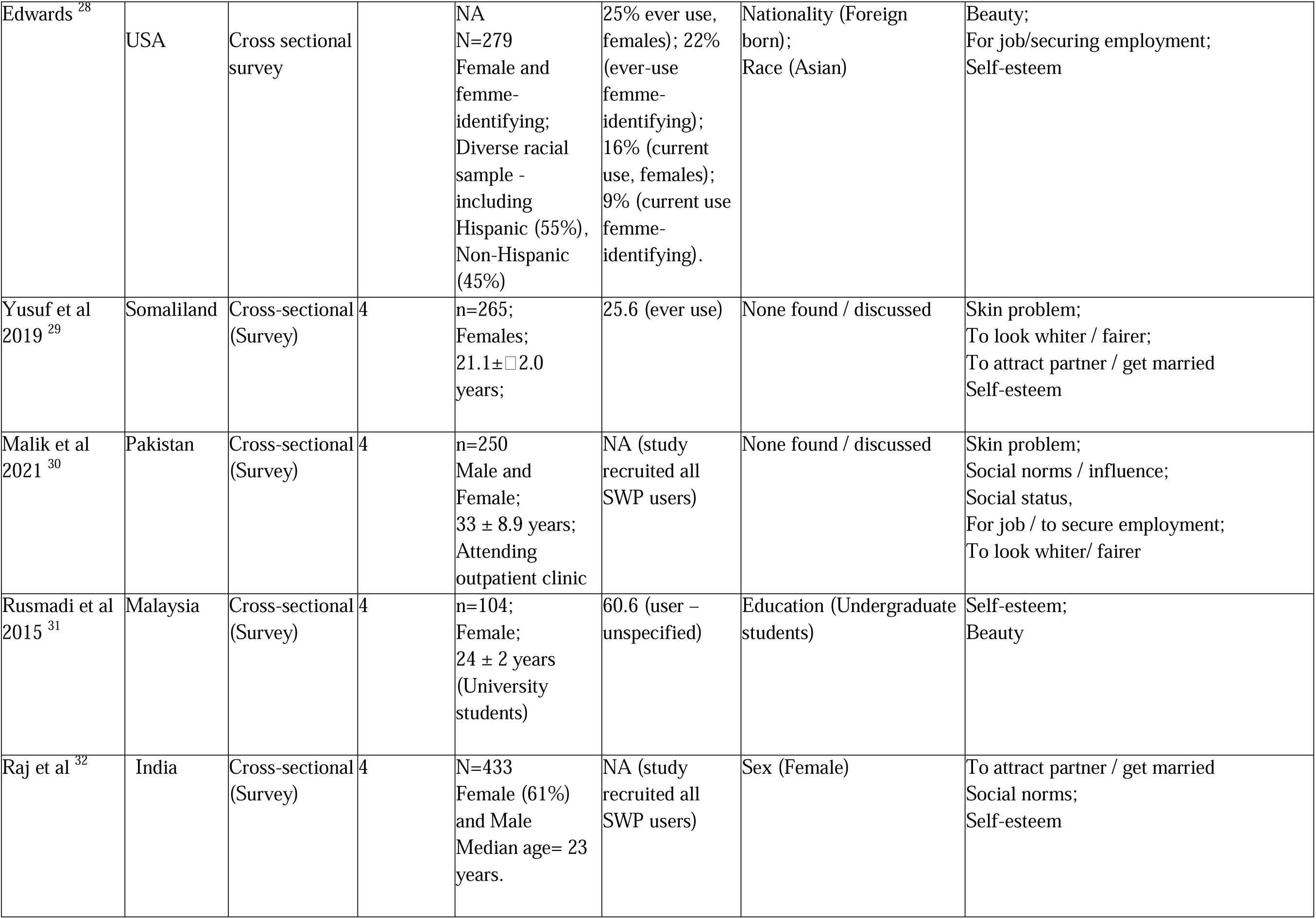
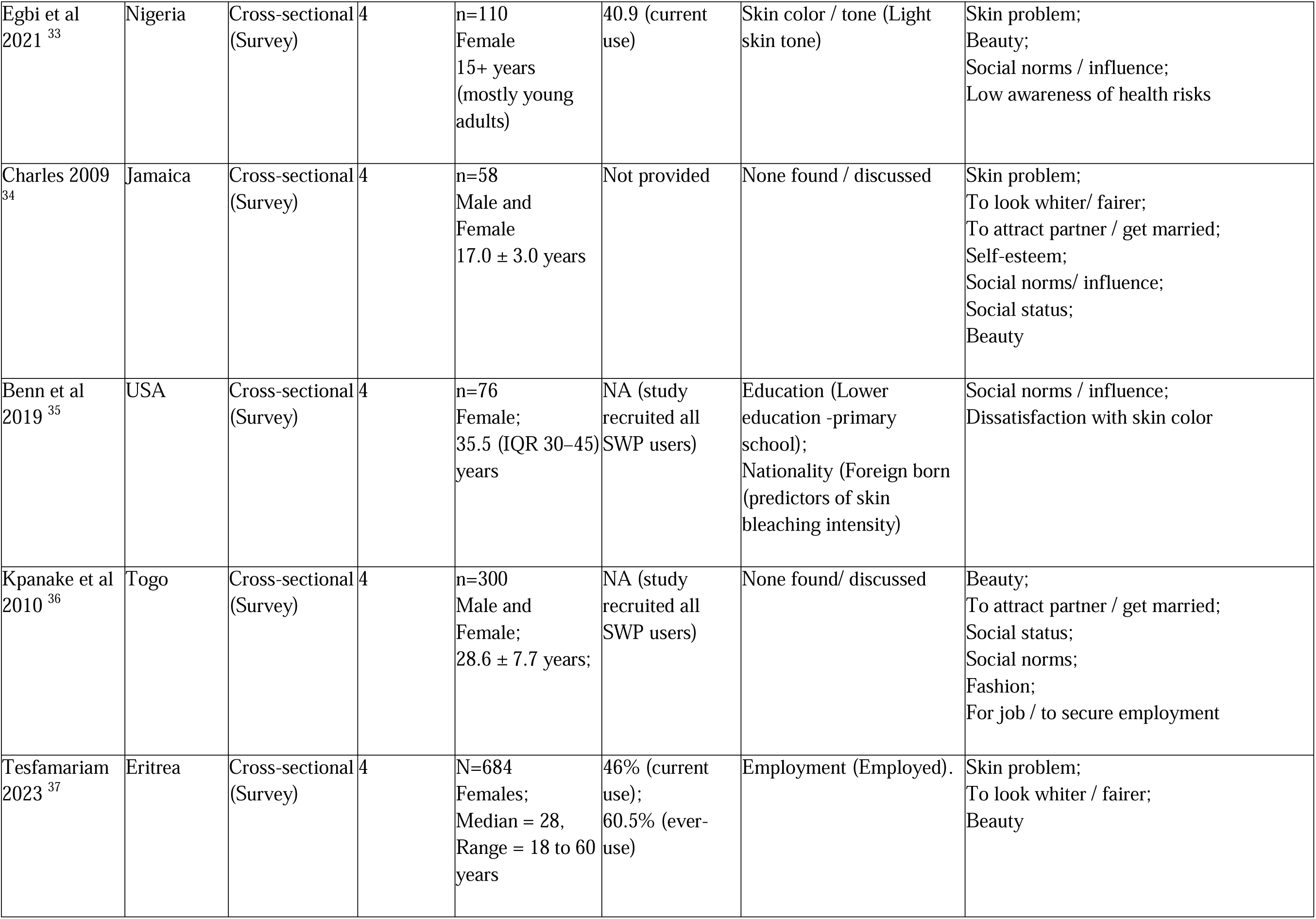
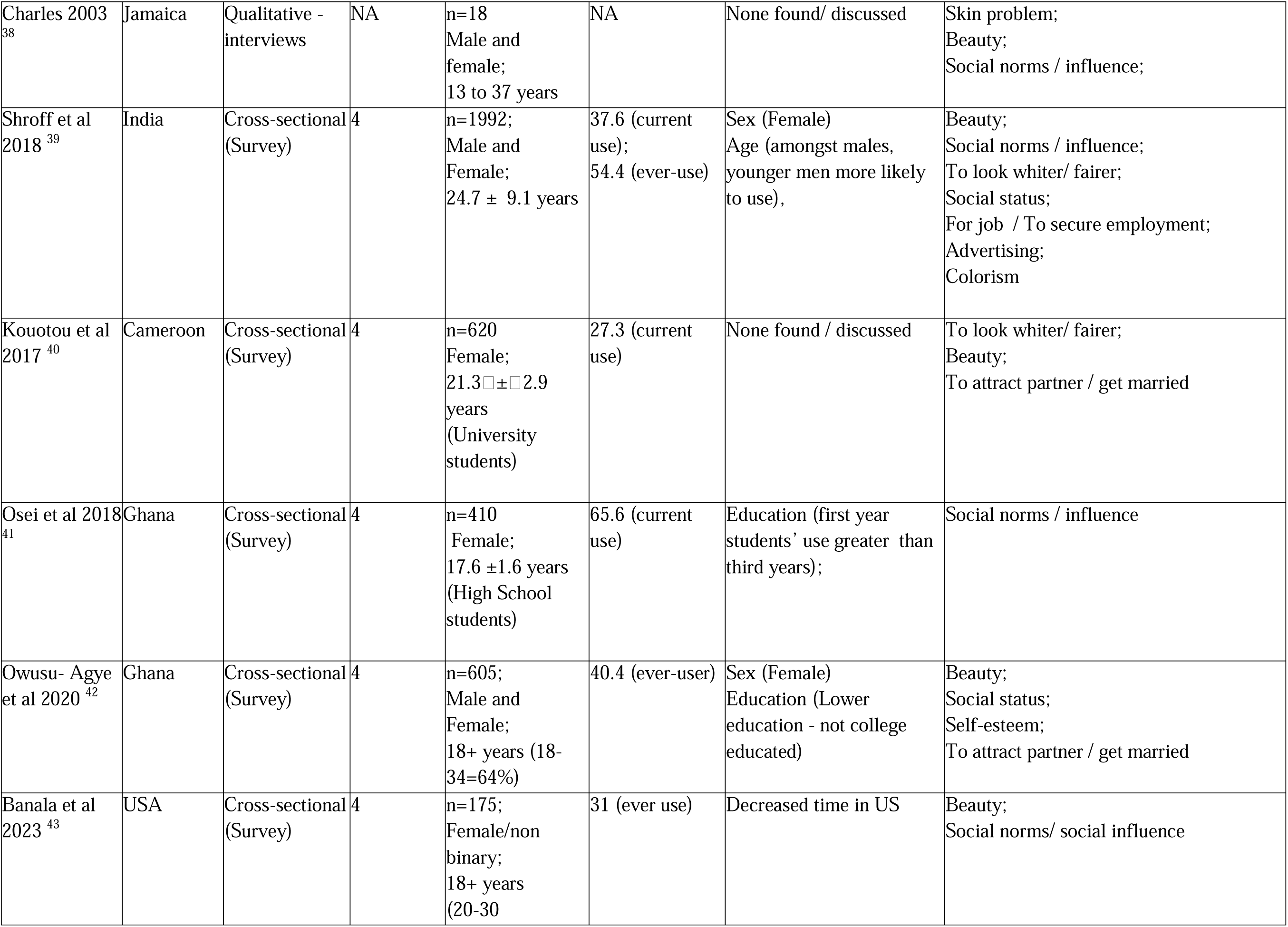
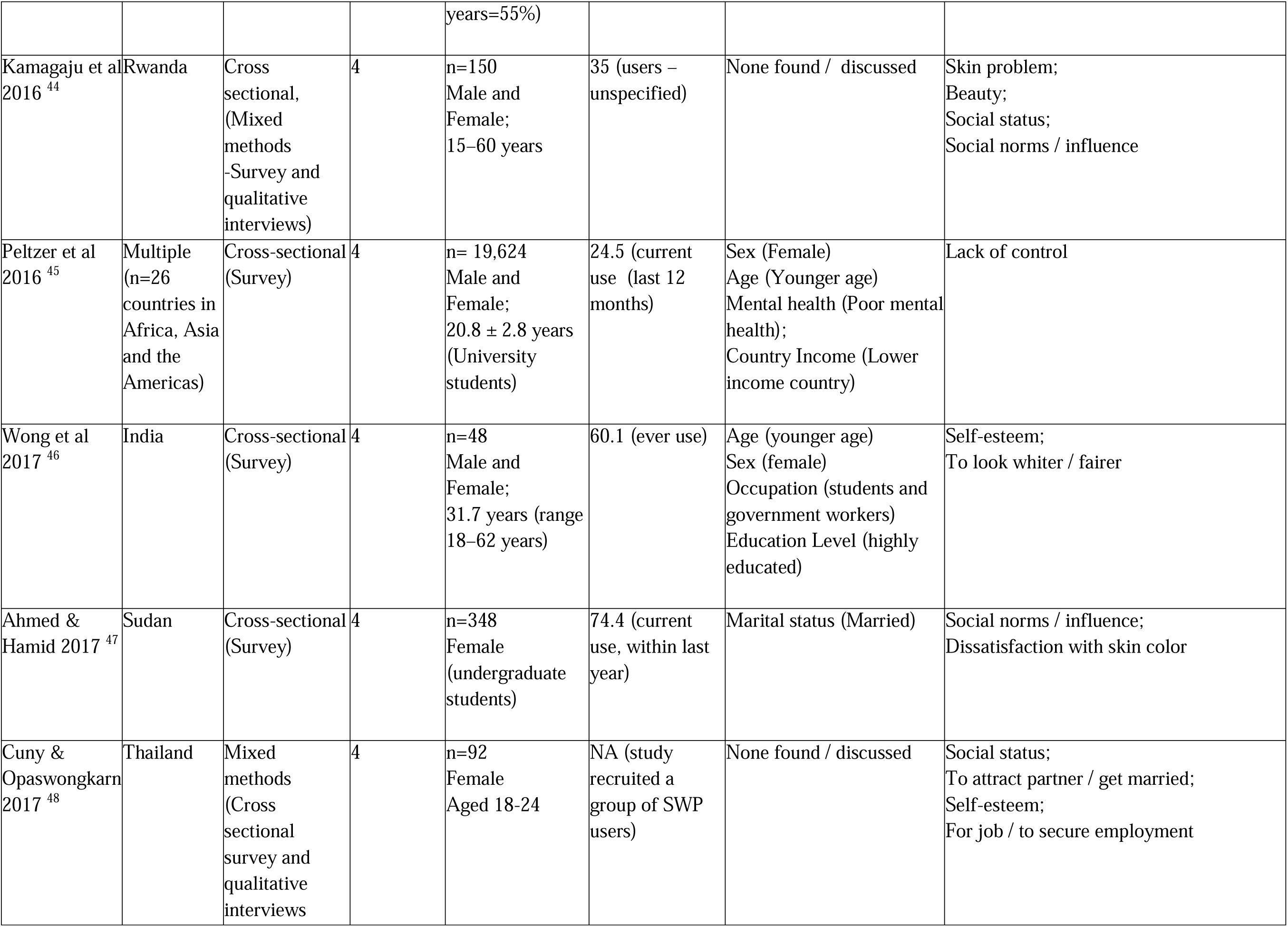
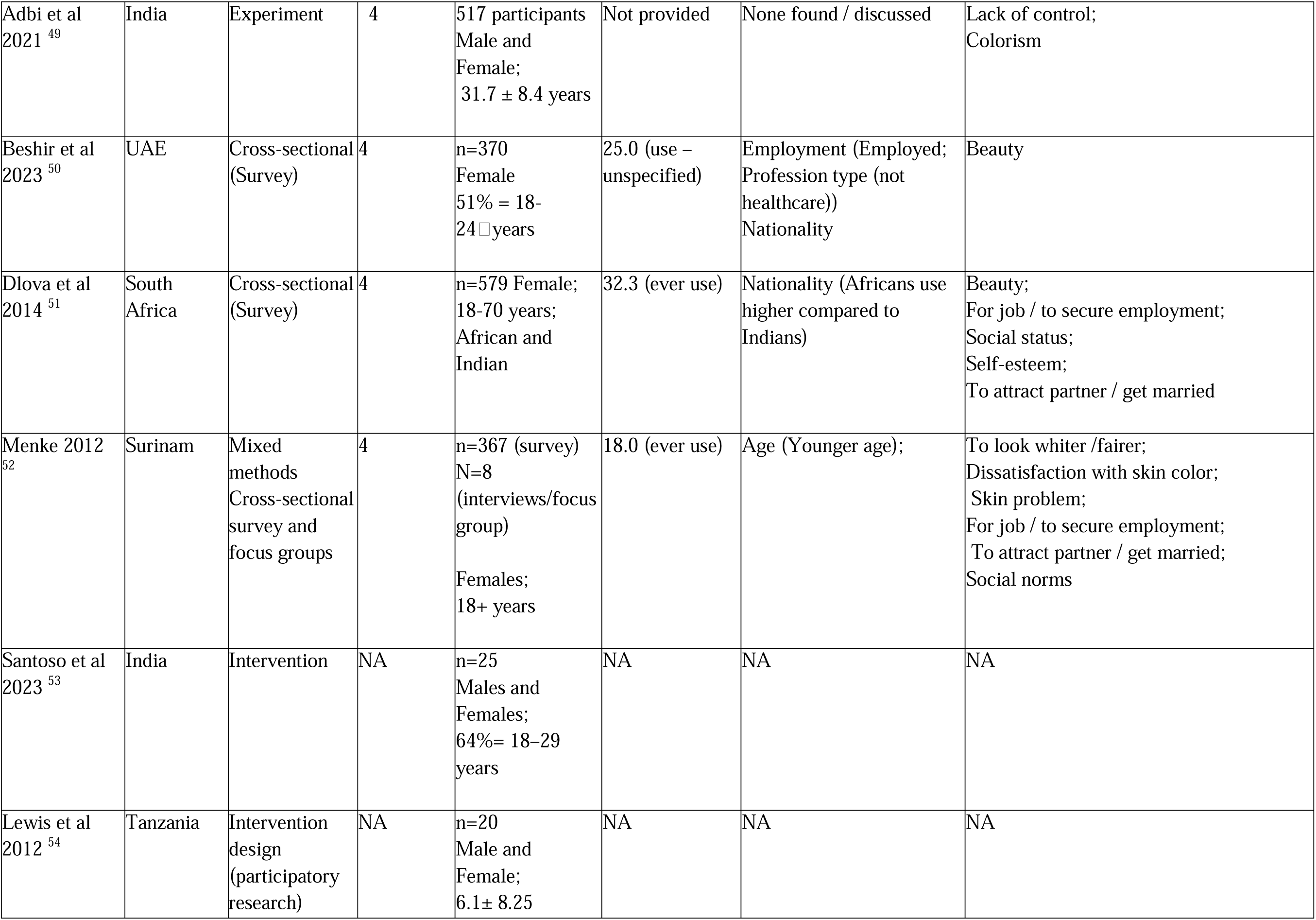
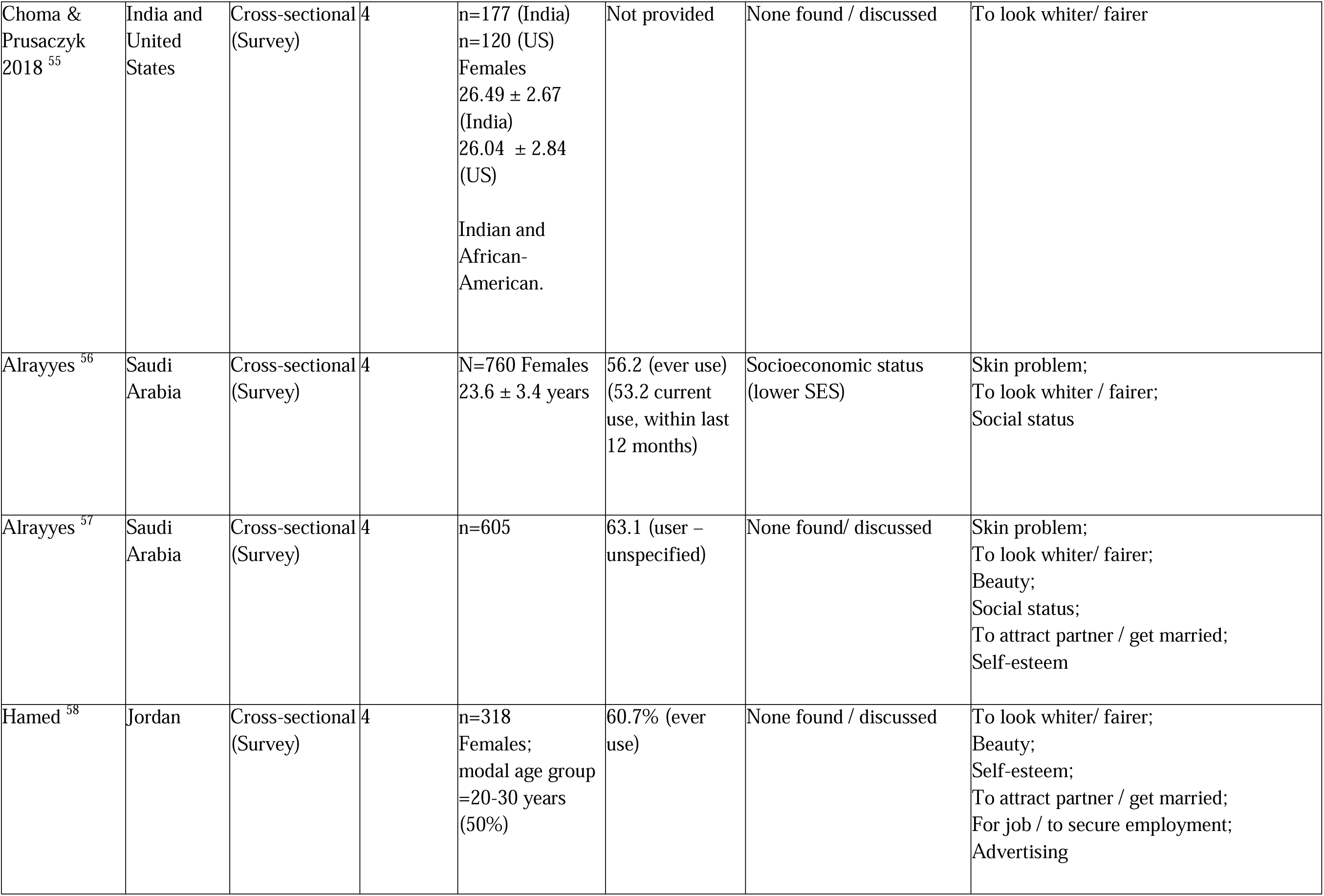
Study Characteristics and Key Themes.

| Ref | Location | Type of Study | OCEM Evidence Level | Participants (N, sex, age, other) | Population prevalence of SLP use (%) | Demographic risk factors (associated with SLP use) | Key Drivers for SLP use |
| --- | --- | --- | --- | --- | --- | --- | --- |
| Darj E, et al 2015 <sup>16</sup> | Sweden | Cross sectional, (Mixed methods -Survey and qualitative interviews) | 4 | n=455<br>Female<br>22 to 50 years (pregnant women); | 2.6 (current use) | Nationality (Foreign born) | Beauty;<br>Social status;<br>Low awareness of health risks;<br>To attract partner / get married |
| Yayehrad et al 2023 <sup>17</sup> | Ethiopia | Cross-sectional (Survey) | 4 | N= 362;<br>Female<br>18-30 years= 32.4%<br>31-45 years= 33.5%<br>46+ = 34.1% | NA (study recruited all SLP users) | None found/ discussed | Skin problem;<br>To look whiter / fairer;<br>Beauty;<br>Self-esteem;<br>Social norms / influence;<br>Low awareness of health risks |
| Bamerdah et al 2023 <sup>18</sup> | Saudi Arabia | Cross-sectional (Survey) | 4 | n=409;<br>Female;<br>18+ years | 35.7 (ever use) | Age (Younger age: 20-30 years);<br>Education (Higher education) | Beauty;<br>Skin problem;<br>Self-esteem;<br>To attract partner / get married;<br>To look whiter / fairer/ preference for lighter skin tone |
| Nyoni-Kachambwa et al 2021 <sup>19</sup> | Zimbabwe | Cross-sectional (Survey) | 4 | n=260<br>Female 18+ years;<br>m=31.69 years | 31.2 (current use) | Employment (Employed);<br>Skin color (Dark skinned);<br>Marital status (Unmarried/ widowed/ | Beauty;<br>Social norms / influence;<br>To attract partner / get married;<br>For job / To secure employment |
|  |  |  |  |  |  | divorced) |  |
| Rahiman et al<br>2021 <sup>20</sup> | South<br>Africa | Cross-sectional<br>(Survey) | 4 | n=401<br>Male and<br>Female<br>(University<br>students);<br>20 ± 2.4 years | 12.0 (users -<br>unspecified) | Location<br>(Urban) | Social norms / influence;<br>Social status;<br>To attract partner / get married;<br>Fashion |
| Daftary et al<br>2023 <sup>21</sup> | USA | Cross-sectional<br>(Survey) | 4 | n=455<br>Male and<br>Female;<br>median= 41 years<br>(IQR=29–54);<br>Black or African<br>American, Asian,<br>multiracial,<br>Hispanic,<br>American Indian<br>or Alaska Native,<br>and other | 21.3 (users -<br>unspecified) | Sex<br>(Female);<br>Skin color<br>(Black);<br>Female;<br>Immigrant status (Second<br>generation) | Skin problem;<br>Beauty;<br>Self-esteem;<br>To attract partner / get married;<br>Colorism |
| Alanzi et al<br>2018 <sup>22</sup> | Saudi<br>Arabia | Cross-sectional<br>(Survey) | 4 | n=531<br>Female;<br>39.9 ± 9.0 years<br>(Attending<br>outpatient<br>clinics) | 43.3 (current<br>use) | Age<br>(Younger age);<br>Education<br>(Higher education);<br>Marital status<br>(Married);<br>Employment<br>(Employed) | Skin problem;<br>To look whiter/ fairer;<br>Beauty;<br>To attract partner / get married<br>Self-esteem;<br>Fashion;<br>Social norms / influence |
| Harper <sup>23</sup> | USA and<br>India | Cross-sectional<br>(Survey) | 4 | n=49 (USA)<br>n=168 (India);<br>Female;<br>29.7 ± 8.5 | Not provided | None found/ discussed | Beauty;<br>Colorism;<br>Dissatisfaction with skin color;<br>To attract partner / get married; |
|  |  |  |  | years;<br>African-American (USA),<br>Indian (India) |  |  | Self-esteem;<br>Social Status |
| Lewis et al<br>2011 <sup>24</sup> | Tanzania | Qualitative -<br>interviews | NA | n=42;<br>Female;<br>m =25.2 years | NA | None found/ discussed | Skin problem;<br>To look whiter/ fairer<br>Beauty;<br>To attract partner / get married;<br>Social norms;<br>Social status |
| Lasco &<br>Hardon 2020<br><sup>25</sup> | Philippines | Qualitative -<br>ethnography | NA | No sample size<br>given; Males | NA | None found/ discussed | Social norms / influence;<br>Fashion;<br>Beauty;<br>For job / To secure employment |
| Peltzer et al<br>2017 <sup>26</sup> | 5 countries<br>in SE Asia | Cross-sectional<br>(Survey) | 4 | n=3259<br>Males and<br>Females;<br>20.5 ± 1.6years, | 30.7 (current<br>use) | Age (Older age amongst<br>female users);<br>Socio-economic status<br>(Low SES amongst female<br>users)<br>Mental health (Poor mental<br>health amongst male and<br>female users) | Beauty;<br>Self-esteem;<br>Low awareness of health risks<br>Social status;<br>For job / to secure employment;<br>To attract partner / get married;<br>Advertising |
| Motlohi et al<br>2023 <sup>27</sup> | Lesotho | Cross-sectional<br>(Survey) | 4 | n=468;<br>Males and<br>Females;<br>Modal age<br>group= 21-25<br>years (23.2%) | 43.7% (current<br>use) | Employment (Manual<br>employment - Factory<br>workers) | Skin problem;<br>Beauty;<br>Low awareness of health risks;<br>Social norms / influence |
| Edwards <sup>28</sup> | USA | Cross sectional survey |  | NA<br>N=279<br>Female and femme-identifying;<br>Diverse racial sample - including Hispanic (55%), Non-Hispanic (45%) | 25% ever use, females); 22% (ever-use femme-identifying); 16% (current use, females); 9% (current use femme-identifying). | Nationality (Foreign born);<br>Race (Asian) | Beauty;<br>For job/securing employment;<br>Self-esteem |
| Yusuf et al 2019 <sup>29</sup> | Somaliland | Cross-sectional (Survey) | 4 | n=265;<br>Females;<br>21.1±2.0 years; | 25.6 (ever use) | None found / discussed | Skin problem;<br>To look whiter / fairer;<br>To attract partner / get married<br>Self-esteem |
| Malik et al 2021 <sup>30</sup> | Pakistan | Cross-sectional (Survey) | 4 | n=250<br>Male and Female;<br>33 ± 8.9 years;<br>Attending outpatient clinic | NA (study recruited all SWP users) | None found / discussed | Skin problem;<br>Social norms / influence;<br>Social status,<br>For job / to secure employment;<br>To look whiter/ fairer |
| Rusmadi et al 2015 <sup>31</sup> | Malaysia | Cross-sectional (Survey) | 4 | n=104;<br>Female;<br>24 ± 2 years (University students) | 60.6 (user – unspecified) | Education (Undergraduate students) | Self-esteem;<br>Beauty |
| Raj et al <sup>32</sup> | India | Cross-sectional (Survey) | 4 | N=433<br>Female (61%) and Male<br>Median age= 23 years. | NA (study recruited all SWP users) | Sex (Female) | To attract partner / get married<br>Social norms;<br>Self-esteem |
| Egbi et al<br>2021 <sup>33</sup> | Nigeria | Cross-sectional<br>(Survey) | 4 | n=110<br>Female<br>15+ years<br>(mostly young<br>adults) | 40.9 (current<br>use) | Skin color / tone (Light<br>skin tone) | Skin problem;<br>Beauty;<br>Social norms / influence;<br>Low awareness of health risks |
| Charles 2009<br><sup>34</sup> | Jamaica | Cross-sectional<br>(Survey) | 4 | n=58<br>Male and<br>Female<br>17.0 ± 3.0 years | Not provided | None found / discussed | Skin problem;<br>To look whiter/ fairer;<br>To attract partner / get married;<br>Self-esteem;<br>Social norms/ influence;<br>Social status;<br>Beauty |
| Benn et al<br>2019 <sup>35</sup> | USA | Cross-sectional<br>(Survey) | 4 | n=76<br>Female;<br>35.5 (IQR 30–45)<br>years | NA (study<br>recruited all<br>SWP users) | Education (Lower<br>education -primary<br>school);<br>Nationality (Foreign born<br>(predictors of skin<br>bleaching intensity) | Social norms / influence;<br>Dissatisfaction with skin color |
| Kpanake et al<br>2010 <sup>36</sup> | Togo | Cross-sectional<br>(Survey) | 4 | n=300<br>Male and<br>Female;<br>28.6 ± 7.7 years; | NA (study<br>recruited all<br>SWP users) | None found/ discussed | Beauty;<br>To attract partner / get married;<br>Social status;<br>Social norms;<br>Fashion;<br>For job / to secure employment |
| Tesfamariam<br>2023 <sup>37</sup> | Eritrea | Cross-sectional<br>(Survey) | 4 | N=684<br>Females;<br>Median = 28,<br>Range = 18 to 60<br>years | 46% (current<br>use);<br>60.5% (ever-use) | Employment (Employed). | Skin problem;<br>To look whiter / fairer;<br>Beauty |
| Charles 2003<sup>38</sup> | Jamaica | Qualitative - interviews | NA | n=18<br>Male and female;<br>13 to 37 years | NA | None found/ discussed | Skin problem;<br>Beauty;<br>Social norms / influence; |
| Shroff et al 2018<sup>39</sup> | India | Cross-sectional (Survey) | 4 | n=1992;<br>Male and Female;<br>24.7 ± 9.1 years | 37.6 (current use);<br>54.4 (ever-use) | Sex (Female)<br>Age (amongst males, younger men more likely to use), | Beauty;<br>Social norms / influence;<br>To look whiter/ fairer;<br>Social status;<br>For job / To secure employment;<br>Advertising;<br>Colorism |
| Kouotou et al 2017<sup>40</sup> | Cameroon | Cross-sectional (Survey) | 4 | n=620<br>Female;<br>21.3 ± 2.9 years<br>(University students) | 27.3 (current use) | None found / discussed | To look whiter/ fairer;<br>Beauty;<br>To attract partner / get married |
| Osei et al 2018<sup>41</sup> | Ghana | Cross-sectional (Survey) | 4 | n=410<br>Female;<br>17.6 ± 1.6 years<br>(High School students) | 65.6 (current use) | Education (first year students' use greater than third years); | Social norms / influence |
| Owusu- Agye et al 2020<sup>42</sup> | Ghana | Cross-sectional (Survey) | 4 | n=605;<br>Male and Female;<br>18+ years (18-34=64%) | 40.4 (ever-user) | Sex (Female)<br>Education (Lower education - not college educated) | Beauty;<br>Social status;<br>Self-esteem;<br>To attract partner / get married |
| Banala et al 2023<sup>43</sup> | USA | Cross-sectional (Survey) | 4 | n=175;<br>Female/non binary;<br>18+ years<br>(20-30 | 31 (ever use) | Decreased time in US | Beauty;<br>Social norms/ social influence |
|  |  |  |  | years=55%) |  |  |  |
| Kamagaju et al<br>2016 <sup>44</sup> | Rwanda | Cross<br>sectional,<br>(Mixed<br>methods<br>-Survey and<br>qualitative<br>interviews) | 4 | n=150<br>Male and<br>Female;<br>15–60 years | 35 (users –<br>unspecified) | None found / discussed | Skin problem;<br>Beauty;<br>Social status;<br>Social norms / influence |
| Peltzer et al<br>2016 <sup>45</sup> | Multiple<br>(n=26<br>countries in<br>Africa, Asia<br>and the<br>Americas) | Cross-sectional<br>(Survey) | 4 | n= 19,624<br>Male and<br>Female;<br>20.8 ± 2.8 years<br>(University<br>students) | 24.5 (current<br>use (last 12<br>months) | Sex (Female)<br>Age (Younger age)<br>Mental health (Poor mental<br>health);<br>Country Income (Lower<br>income country) | Lack of control |
| Wong et al<br>2017 <sup>46</sup> | India | Cross-sectional<br>(Survey) | 4 | n=48<br>Male and<br>Female;<br>31.7 years (range<br>18–62 years) | 60.1 (ever use) | Age (younger age)<br>Sex (female)<br>Occupation (students and<br>government workers)<br>Education Level (highly<br>educated) | Self-esteem;<br>To look whiter / fairer |
| Ahmed &<br>Hamid 2017 <sup>47</sup> | Sudan | Cross-sectional<br>(Survey) | 4 | n=348<br>Female<br>(undergraduate<br>students) | 74.4 (current<br>use, within last<br>year) | Marital status (Married) | Social norms / influence;<br>Dissatisfaction with skin color |
| Cuny &<br>Opaswongkarn<br>2017 <sup>48</sup> | Thailand | Mixed<br>methods<br>(Cross<br>sectional<br>survey and<br>qualitative<br>interviews | 4 | n=92<br>Female<br>Aged 18-24 | NA (study<br>recruited a<br>group of SWP<br>users) | None found / discussed | Social status;<br>To attract partner / get married;<br>Self-esteem;<br>For job / to secure employment |
| Adbi et al<br>2021 <sup>49</sup> | India | Experiment | 4 | 517 participants<br>Male and<br>Female;<br>31.7 ± 8.4 years | Not provided | None found / discussed | Lack of control;<br>Colorism |
| Beshir et al<br>2023 <sup>50</sup> | UAE | Cross-sectional<br>(Survey) | 4 | n=370<br>Female<br>51% = 18-24□years | 25.0 (use –<br>unspecified) | Employment (Employed;<br>Profession type (not<br>healthcare))<br>Nationality | Beauty |
| Dlova et al<br>2014 <sup>51</sup> | South<br>Africa | Cross-sectional<br>(Survey) | 4 | n=579 Female;<br>18-70 years;<br>African and<br>Indian | 32.3 (ever use) | Nationality (Africans use<br>higher compared to<br>Indians) | Beauty;<br>For job / to secure employment;<br>Social status;<br>Self-esteem;<br>To attract partner / get married |
| Menke 2012<br><sup>52</sup> | Surinam | Mixed<br>methods<br>Cross-sectional<br>survey and<br>focus groups | 4 | n=367 (survey)<br>N=8<br>(interviews/focus<br>group)<br>Females;<br>18+ years | 18.0 (ever use) | Age (Younger age); | To look whiter /fairer;<br>Dissatisfaction with skin color;<br>Skin problem;<br>For job / to secure employment;<br>To attract partner / get married;<br>Social norms |
| Santoso et al<br>2023 <sup>53</sup> | India | Intervention | NA | n=25<br>Males and<br>Females;<br>64%= 18–29<br>years | NA | NA | NA |
| Lewis et al<br>2012 <sup>54</sup> | Tanzania | Intervention<br>design<br>(participatory<br>research) | NA | n=20<br>Male and<br>Female;<br>6.1± 8.25 | NA | NA | NA |
| Choma & Prusaczyk 2018 <sup>55</sup> | India and United States | Cross-sectional (Survey) | 4 | n=177 (India)<br>n=120 (US)<br>Females<br>26.49 ± 2.67 (India)<br>26.04 ± 2.84 (US)<br>Indian and African-American. | Not provided | None found / discussed | To look whiter/ fairer |
| Alrayyes <sup>56</sup> | Saudi Arabia | Cross-sectional (Survey) | 4 | N=760 Females<br>23.6 ± 3.4 years | 56.2 (ever use)<br>(53.2 current use, within last 12 months) | Socioeconomic status (lower SES) | Skin problem;<br>To look whiter / fairer;<br>Social status |
| Alrayyes <sup>57</sup> | Saudi Arabia | Cross-sectional (Survey) | 4 | n=605 | 63.1 (user – unspecified) | None found/ discussed | Skin problem;<br>To look whiter/ fairer;<br>Beauty;<br>Social status;<br>To attract partner / get married;<br>Self-esteem |
| Hamed <sup>58</sup> | Jordan | Cross-sectional (Survey) | 4 | n=318 Females;<br>modal age group =20-30 years (50%) | 60.7% (ever use) | None found / discussed | To look whiter/ fairer;<br>Beauty;<br>Self-esteem;<br>To attract partner / get married;<br>For job / to secure employment;<br>Advertising |

**Table 2:** Behavioral factors under the three domains of the WHO principles, definitions and frequency and countries from which the findings derive.

| Domain/Theme □ | Definition □ | Frequency and citations □ | Countries □ |
| --- | --- | --- | --- |
| 1. Psychological and cognitive factors: Factors related to how people think and make decisions, for example cognition, knowledge, motivation, emotion, biases and heuristics, conscious and unconscious. □ |  |  |  |
| 1.1. Beauty□□ | The aesthetic use of SLPs to make the user feel or be seen as more attractive or more beautiful.□ | 26 ( <sup>16 17 19 21 22 23 24 25 26 27 28 31 33 34 36 37 38 39</sup><br><sup>40 42 43 44 50 51 57 58</sup> )□ | Sweden, Ethiopia, Zimbabwe, USA (4), Saudi Arabia (2), India (2), Tanzania, Philippines, Lesotho, Malaysia (2), Nigeria, Jamaica (2), Togo, Eritrea, Cameroon, Ghana, Rwanda, UAE, South Africa, Jordan, Indonesia, Myanmar, Thailand, Vietnam□□ |
| 1.2. Self-efficacy□□ | The use of SLPs to feel more confident or to enhance their feelings of self-worth.□ | 17 ( <sup>17 18 21 22 23 26 28 29 31 32 34 42 46 48 51 57 58</sup> )□ | Ethiopia, Saudi Arabia (3), USA (3), India (3), Tanzania, Somaliland, Malaysia (2), Jamaica, Ghana, Thailand (2), South Africa, Jordan, Indonesia, Myanmar, Vietnam□ |
| 1.3. Attract a partner or get married□ | The use of SLPs to feel more attractive to a partner or with the goal of getting married.□ | 20 ( <sup>16 18 19 20 21 22 23 24 26 29 32 34 36 40 42 48 51 52</sup><br><sup>57 58</sup> )□ | Sweden, Saudi Arabia (3), Zimbabwe, South Africa (2), USA (2), India (2), Tanzania, Somaliland, Jamaica, Togo, Cameroon, Ghana, Thailand (2), Surinam, Jordan, Indonesia, Malaysia, Myanmar, Vietnam□ |
| 1.4. Dissatisfaction with skin tone□□ | The use of SLPs due to being unhappy with the color or tone of skin and wanting to change skin color or tone.□ | 4 ( <sup>23 35 47 52</sup> )□ | USA (2), India, Sudan, Surinam□ |
| 1.5. Low awareness of risks of skin whitening□□ | Lack of knowledge or awareness of the health risks, side effects or harmful ingredients of SLPs as a barrier to abstaining from their use.□□ | 5 ( <sup>16 17 26 27 33</sup> )□ | Sweden, Ethiopia, Indonesia, Malaysia, Myanmar, Thailand, Vietnam□ |
| 1.7. Look whiter or fairer□ | The use of SLPs to look 'whiter' or 'fairer', including specifically to look 'more European'.□ | 16 ( <sup>17 18 22 24 29 30 34 37 39 40 46 52 55 56 57 58</sup> )□ | Ethiopia, Saudi Arabia (4), Tanzania, Somaliland, Pakistan, Jamaica, Eritrea, India (3), Cameroon, Surinam, USA, Jordan |
| 2. Social and cultural factors: Factors related to the social and cultural contexts within which behaviours take place, for example social norms and practices, local identity and values, social support systems and cultural commitments.□ |  |  |  |
| 2.1. Social status□□ | The use of SLPs to enhance one's social status, related to the social perception or bias that lighter skin is associated with higher social status.□ | 15 ( <sup>16 20 23 24 26 30 34 36 39 42 44 48 51 56 57</sup> )□ | Sweden, South Africa (2), USA, India (2), Tanzania, Pakistan, Jamaica, Togo, Ghana, Rwanda, Thailand (2), |
|  |  |  | Saudi Arabia (2), Indonesia, Malaysia, Myanmar, Vietnam□ |
| 2.2. Social norms or influence□ | The modelling and/or encouragement of SLP use by significant others, including family, friends or other social networks.□ | 20 (<sup>17 19 20 22 24 25 27 30 32 33 34 35 36 38 39 41 43 44 47 52</sup>)□ | Ethiopia, Zimbabwe, South Africa, Saudi Arabia, Tanzania,□ Philippines, Lesotho, Pakistan, India (2), Nigeria (2), USA (2), Togo, Jamaica, Ghana, Rwanda,□ Sudan, Surinam□ |
| 2.3. Colorism□ | The privileging of and preference for lighter skin within ethnic and racial groups as a cultural factor or pressure for SLP use.□ | 3 (<sup>21 23 49</sup>)□ | USA (2), India (2).□ |
| 2.4. For a job or to secure employment□ | The use of SLPs due to the perception that□ having lighter skin conveys a more 'professional' look or will aid employability; linked to social inequalities around skin colour and hiring practices.□ | 9 (<sup>19 25 26 30 36 39 48 52 58</sup>)□ | Zimbabwe, Philippines, Pakistan, Togo, India, Thailand, Surinam, Jordan, Indonesia, Malaysia, Myanmar, Thailand, Vietnam□□ |
| 3. Environmental factors: Factors external to the individual which could influence the behaviour, including physical or social environment or setting.□ |  |  |  |
| 3.1. Advertising□ | The influence of advertising and promotion of SLPs – including via product packaging, TV and other traditional advertising and via social media.□ | 3 (<sup>26 39 58</sup>)□ | India, Jordan, Indonesia, Malaysia, Myanmar, Thailand, Vietnam□ |
| 3.2. Fashion□ | The role of fashion (as a concept and industry) in promoting the use of SLPs by portraying lighter skin achieved via SLP use as fashionable, trendy or desirable.□ | 4 (<sup>20 22 25 36</sup>)□ | South Africa, Saudi Arabia, Philippines, Togo.□ |

### (a) What are the main behavioural factors (psychological and cognitive, social and cultural, environmental) behind the practice of skin whitening?

This review focuses on the cosmetic use of skin whitening - rather than the use of clinically prescribed SLPs for specific medical conditions such as melasma. However, it is important to note that a commonly stated reason for the use of skin whitening products was to alleviate skin related issues; that is self-defined problems such as ‘to remove pimples or rashes’ or to ‘remove facial blemishes’ (n=16).^17,18,21,22,24,27,29,30,33,34,37,38,44,52,56,57^ In some instances, this is a key reason for the initial onset of use, with other factors helping to explain continued use. Overall, this review found twelve main themes related to the most common factors explaining the cosmetic practice of skin whitening: *Beauty*; *Self-esteem; To attract a partner or get married; Dissatisfaction with skin tone; Low awareness of health risks; Whiteness; Social status; Social norms or social influence; For a job or to secure employment; Colorism; Influence of Advertising;* and *Fashion*.

Behaviors such as cosmetic skin whitening are the product of a complex interplay between psychological and cognitive, social and cultural, and environmental factors. However, we discuss key themes for clarity under their primary designation (e.g. social status as primarily a social factor).

#### Psychological and cognitive factors

*Beauty or attractiveness* was noted in 26 studies. ^16,17,19,21,22,23,24,25,26,27,28,31,33,34,36,37,38,39,40,42,43,44,50,51,57,58^ The practice of skin whitening was seen as something that would make the user feel or be seen as more attractive or more beautiful. This theme was related to social stereotypes and notions of beauty - and perceptions of fairer skin as more ‘attractive’. 17 studies identified *self-esteem* as a reason why participants were more likely to use skin whitening products.^17,18,21,22,23,26,28,29,31,32,34,42,46,48,51,57,58^ Specifically, lighter skin was seen to enhance feelings of self confidence in users. Whitening skin for an existing partner, or to *attract a partner or to get married* was found to be a driver of use in 20 studies ^16,18,19,20,21,22,23,24,26,29,32,34,36,40,42,48,51,52,57,58^. In particular, studies discussed how female participants used skin whitening products for a husband, male partner or in order to be attractive to men, reflecting the gendered nature of this practice in many countries and contexts (although some studies found that skin whitening was not significantly driven by wanting to be attractive for men or a partner ^33,46^). Four studies found that participants’ feelings of dissatisfaction with their skin tone was a key factor ^23,35,47,52^. This related to the theme of generally wanting to *appear whiter*, fairer or, in some studies, more ‘European-looking’ (n=16). ^17,18,22,24,29,30,34,37,39,40,46,52,55,56,57,58^ Finally, a lack of knowledge or awareness of the health risks associated with skin whitening was a significant factor in 5 studies, ^16,17,26,27,33^ although one study found that skin whitening use occurred despite awareness of side effects or health risks.^46^

#### Social and cultural factors

The practice of skin whitening to enhance the perception of one’s *social status* was found in 15 studies.^16,20,23,24,26,30,34,36,39,42,44,48,51,56,57^ Specifically, the association was made between lighter or fairer looking skin and the aspiration to look or be seen as having higher social status. The role of *social norms or social influence* was a key theme in 20 studies.^17,19,20,22,24,25,27,30,32,33,34,35,36,38,39,41,43,44,47,52^ Studies discussing this theme emphasized the role that social norms play in shaping individual perceptions around the desire to have lighter skin or that fairer skin is socially advantageous. The practice of skin whitening to *for a job or to secure employmen*t was found in 9 studies.^19,25,26,30,36,39,48,52,58^ These studies discussed how having lighter skin was seen as being more professional, or specifically as an advantage to securing a job or to advancing one’s career.

A cultural driver identified was the influence of colorism. Colorism is the privileging of and preference for lighter skin within ethnic and racial groups.^23^ Three studies explicitly studied the association between colorism and skin whitening product use.^21,23,49^ One study, from the United States found that colorism scores (i.e. perceptions that lighter skin is more desirable and advantageous) were significantly higher amongst those who used skin whitening products, compared to those who didn’t use them.^21^ A second study with African-American women found that skin whitening behavior was associated with the internalization of white beauty ideals.^23^ Another study, from India, explored how culturally-ingrained colorism shaped individual level feelings of disempowerment to predict women’s preference for stronger and riskier skin lightening products.^49^

#### Environmental factors

The significant role that advertising and media play in encouraging skin lightening was mentioned in three studies.^26,39,58^ This theme discussed the influence of advertising and promotion of skin whitening products (including commercially and via social media). For example, one study found that media, TV and advertising were among the most common prompts for skin lightening use.^39^ These studies did not however observe or study purchasing behavior. Relatedly, the influence of the fashion or ‘fad’ of skin whitening products was discussed in four studies.^20,22,25,36^ For example, one ethnographic study explored how the role of product advertising and changing notions of fashion and masculinity amongst young men in the Philippines has contributed to the growing popularity of skin whitening amongst this group.^25^

### (b) Which groups and characteristics are associated with the practice of skin whitening?

Prevalence of use ranged from 2%-74% (although studies varied in terms of whether they measured current or recent use and ever/lifetime use, and in terms of how the latter was defined). The evidence on the demographic characteristics associated with the practice of skin whitening revealed mixed results, but analysis of included studies identified the following key correlates: (1) *Sex*; (2) *Age* (3) *Education (4) Employment*. Studies identified a number of other demographic correlates, for example, being employed and being married, although these did not feature commonly enough to be classed as themes.

#### Gender

In keeping with an existing review of the prevalence of skin whitening,^9^ the studies included in this review also suggest that skin whitening is a unisex practice, but more common amongst females. Although a number of studies exclusively recruited female participants, where both males and females were studied, use of skin whitening products was observed amongst both genders, although five studies found that being female significantly predicted skin whitening product use, or more intensive product use.^21,26,39,42,45^

#### Age

In general, younger ages are more likely to practice skin whitening. Although most studies were limited in terms of sample selection - being non-representative, non-probability samples (e.g. with some being opportunity samples of higher education students), some did include general population samples, including a range of ages. Where age was studied as a potential predictor of skin whitening, the practice was seen to be more common amongst younger adults in 6 studies.^18,22,39,45,46,52^

#### Education

Three studies found that practice was higher amongst those with lower education,^35,42,46^ whereas two found it was associated with those with higher education (university degrees).^18,22^ This may be country and culturally specific - for example, two studies from Saudi Arabia found an association between skin whitening and higher formal education, ^18,22^ whereas some studies in other countries (e.g. Ghana and the USA) found an association between low formal educational status and higher skin whitening prevalence.^35,42,46^ This relationship may be mediated by the strength of certain factors (e.g. social status) relative to others (e.g. lack of awareness of health risks), which may differ across countries and populations. However, this question was not specifically addressed by the included studies, which as noted were almost exclusively single-country studies.

### (c) What interventions are designed to address the practice of skin whitening and what, if any, evidence is there of their effectiveness?

Only one study was identified which reported results of an implemented intervention specifically designed to address the practice of skin whitening.^53^ This study, from India described an online brief 3-week online-course, using a ‘storytelling’ and social change approach to public health communication, designed to increase participants’ knowledge of and concern for the seriousness of skin-shade discrimination and the use of skin-lightening products. Post-course results revealed a significant increase in participants’ knowledge of skin shade discrimination and knowledge of use of skin whitening products.^53^ However, the study was limited in so far as it did not include a control group, had few participants (n=25) and did not measure whether the intervention affected participants’ intentions to, or actual practice of, skin whitening. One other study, from Tanzania, described a focus group, participatory-based piece of research which aimed to design components of an intervention to address the problem of skin whitening in the community.^54^ It found that key components should include didactic education, governmental action, and educational media.^54^ However, the study did not include the implementation, or empirical evaluation of an intervention based on these principles.

## 4. Discussion

The health risks associated with skin whitening are well documented,^2^ and there is a growing appreciation that underlying factors driving skin lightening must be understood and addressed to prevent and stop this harmful practice. Cosmetic skin whitening has long standing roots in global racism and colonial history.^13^ Skin whitening needs therefore to be studied and understood in relation to its broad historical and cultural context, where - although racism manifests differently in different countries and cultures, white skin and lighter skin tones have been privileged and associated with higher status and power. However, while colorism and racism are tackled as part of long-term complex strategies, public health interventions can help to understand how to reduce the use of harmful products in the short-term. This systematic review is to our knowledge the first to synthesize evidence on the behavioral factors explaining the use of SLPs and an attempt of describing and explaining human behavior in this space so to design effective public health interventions aimed The review found 43 studies drawing out 12 themes representing the psychological and cognitive, social and cultural, and environmental factors leading to skin whitening practice: *Beauty*; *Self-esteem*; *To attract a partner or get married; men or getting married; Dissatisfaction with skin tone; Low awareness of risks of skin whitening*; *To look whiter or fairer; Social status*; *Social norms or influence; For a job or to seek employment; Colorism; Influence of Advertising; and Fashion*.

A strength of the body of literature included in this review is that it was global in scope, covering study populations from a diverse range of countries across different regions. However, many of the prominent themes identified were common across countries. For example, the association of skin whitening, and skin fairness, with beauty and attractiveness was found in studies in diverse contexts including Southeast Asia, Africa and North America. It should be noted that a limitation of this review is that, although we classified studies using OCEBM levels of evidence, no formal study quality or risk of bias assessment was conducted. A limitation of the literature is the lack of cohort or case-control studies. Future research should also include longitudinal studies which follow skin whitening practices over time. Also, multi-country studies with representative samples (e.g. including diverse age ranges, SES and education levels) would enable more direct comparisons across countries and sub-populations.

A further limitation of the literature is that most studies did not use theoretical frameworks to elucidate the reasons behind SLP use. Additionally, the complex interplay of psychological and cognitive, social and cultural, and environmental factors was often not explored. Despite the preponderance of studies looking at individual level psychological factors (e.g. perceptions of self-esteem), far fewer studies focused on the role of social and cultural (e.g. colorism) and environmental factors (e.g. advertising or access to). Fewer still attempted to explore how environmental factors like advertising shape, and are shaped by, individual-level factors like perceptions of beauty. For example, aside from a small number of studies,^21,23,49^ more research is needed on the role of colorism and the societal privileging of lighter skin tones on psychological drivers of skin whitening product use including users’ self-esteem and internalized dissatisfaction with their natural skin tone.^59^ The intersectionality of race and gender inequalities is also an important avenue for future research exploring how the power and privilege associated with white skin or light skin tones particularly shapes beauty ideals, particularly affecting women.^8, 59,60^ Although many themes were common across multiple countries and regions, it is also important that research acknowledges the particular racial histories and cultural context of specific countries and populations, including those with links to colonialism.^59,61^ Future research should also look to more intentionally incorporate behavior change theories to apply the research to concrete interventions and strategies aimed at addressing use of harmful products; this would allow for a fuller understanding of the factors behind skin whitening, as identified in this review. For instance, social norms are key predictors of behavior in Theory of Planned Behavior (TPB) and Theory of Normative Social Behavior (TNSB).^62,63^ Self-efficacy is a key predictor in Social Cognitive Theory.^64^ Future research should endeavor to explore skin whitening attitudes, knowledge, intentions and behavior in relation to well-validated theoretical models. Addressing health harming practices requires addressing the underlying psychological and cognitive, social and cultural and environmental factors with targeted behavioral interventions and behavior change techniques. Behavioral theory can help practitioners to describe, explain and predict behaviors of interest. Such interventions should focus on the factors influencing or inhibiting behavior across the different types of factors. Theoretical models, such as Social Cognitive Theory, the COM-B (Capabilities, Opportunities, Motivations and Behavior) model and the socio-ecological model, attempt to acknowledge and account for the complex interplay between individual psychological, attitudinal and cognitive factors and wider social, cultural and environmental factors.^64,65,66^ For example, individual perceptions of dissatisfaction with darker skin tones, or perceptions of fairer skin as being more attractive, cannot be fully understood independently of social norms or stereotypes of beauty, cultural notions of colorism, or the potential exacerbation or exploitation of such perceptions by commercial interests of skin whitening product manufacturers and marketers. Frameworks such as the Behavior Change Wheel (BCW)^65^ can help inform the design of targeted interventions, including education, such as public health campaigns; persuasion or modelling to shift motivation; or regulation, for example through warning labels on products. Social norms could be addressed through social support or persuasion-based interventions, involving trusted sources of information - as identified by the users- and direct influencers, such as family members and peers. Regulation to remove or restrict the sale of harmful products or ingredients would also reduce harm and has little downside from a public health perspective.

The aim of the review has been to provide an empirical basis for the subsequent design of strategic and targeted behavioral interventions based on the ‘diagnosis’ of behavioral factors, including the enablers and barriers, which help explain the practice of cosmetic skin whitening. There is a need for public health practitioners to develop and evaluate tools which can more fully understand the processes by which people start and maintain use of SLPs for cosmetic reasons e.g. the ‘user journey’. Future interventions should be rigorously informed by a theory-based diagnosis of the factors influencing behaviors and ideally evaluated via high-quality randomized controlled trials, grounded in behavioral science theory,^67^ and can be informed by the psychological and cognitive, social and cultural, environmental factors of skin whitening identified in this review.

## Data Availability

The study is a systematic review and contains no primary data.

## Footnotes

1 Although racism persists and affects persons of color in many countries and contexts globally, it has not automatically created the need for SLP use, but has also led to other ways of acceptance, for example being in social settings traditionally reserved for white people. This is outside the scope of this review, but it is important to reiterate that skin color-based discrimination has many forms and consequences outside of the context of skin whitening.

## Conflict of Interest Disclosures

### Author Contributions

Concept and design: Williams, Webb, Jardan, Onyon, Altieri.

Acquisition, analysis, or interpretation of data: Williams, Webb, Lohiniva.

Drafting of the manuscript: Williams.

Critical revision of the manuscript for important intellectual content: All authors.

Administrative, technical, or material support: All authors

Supervision: Altieri.

### Conflict of Interest

No conflicts of interest are reported for this study.

### Funding/Support

This study is part of a project funded by the Global Environment Facility (GEF), implemented by the UN Environment Programme (UNEP) and co-executed by the World Health Organization (WHO) and the Biodiversity Research Organization (BRI) with targeted technical assistance by the UNEP Global Mercury Partnership.

### Role of the Funder/Sponsor

The funding source had no role in the design and conduct of the study; collection, management, analysis, and interpretation of the data; preparation, review, or approval of the manuscript; and decision to submit the manuscript for publication.

### Disclaimers

The content is solely the responsibility of the authors and does not necessarily represent the official views of the World Health Organization.

